# Social mobility and long-term episodic memory in Britain

**DOI:** 10.64898/2026.04.12.26350709

**Authors:** Gindo Tampubolon

## Abstract

Population ageing increases the importance of cognitive capacity for making decisions about retirement and living independently beyond it. We tested whether post-war educational expansion and working-life social mobility eliminate the association between social class of origin and cognition in early old age using the 1958 National Child Development Study. Two outcomes were analysed at age 62: standard episodic memory (immediate + delayed word recall) and long-term episodic memory, capturing accurate half-century recall of childhood household facts (rooms and people at age 11 validated against mothers’ responses). Social mobility trajectories derived in prior work were classified into predominantly manual versus non-manual class trajectories. Models were estimated separately for women and men across three specifications: (i) social origin and controls, (ii) adding social mobility, and (iii) adding weighting to address healthy survivor bias. Education was consistently associated with both outcomes. For long-term episodic memory, social origin gradients were clearer than for short-term episodic memory, with men from service/professional origins showing a 13 percentage-point higher probability of accurate half-century recall than men from manual origins. These findings indicate that education expansion and working-life social mobility failed to release the grip of social origin on long-term episodic memory.

## Introduction

Population ageing is a defining challenge for Britain and other high- and middle-income countries.^1–3^ Longevity gains combined with low fertility have pushed population structures upward, increasing the number of people living for extended stage beyond midlife. This demographic transition has broadened the policy challenge from dependency ratio to capacity: whether older adults can sustain the physical, psychological and cognitive capacities needed to live well and to navigate complex institutional environments.^3–5^

Cognitive function is therefore central to ageing research.^6–8^ Cognitive function supports everyday functioning and shapes the ability to evaluate and act on information. Among cognitive domains, episodic memory (a sum of immediate and delayed recall) is clinically relevant and practically salient because it supports planning and decision-making.^9,10^ Cognitive ageing research faces two recurring threats to inference. First, longitudinal studies of older adults are susceptible to healthy survivor bias and non-random attrition.^6^ Individuals who are frailer, poorer or cognitively weaker are more likely to lost to attrition, and failure to account for this process can distort estimated trajectories and risk-factor associations. Second, many life-course investigations rely on retrospective accounts of childhood conditions collected in old age. Though valuable, retrospective reports are often error-laced and can be biased.^7,11^ These challenges of healthy survivor bias and recall error critically determine what can be safely claimed about cognitive ageing and its social determinants.

Evidence is mounting, including in this journal, to show that childhood circumstances continue to influence health and functioning in older age across many countries and diverse outcomes including episodic memory, Alzheimer’s disease-related dementia, gait speed, grip strength, depression, inflammation, frailty and multimorbidity.^7,12–16^ This life course shaping of health thesis implies that early-life social conditions can leave durable marks that persist into late life.^17–19^ One candidate pathway is that childhood conditions channel individuals into adult socioeconomic positions and exposures; another is biological embedding, including epigenetic processes (e.g., methylation), through which childhood conditions may go ‘under-the-skin’ and influence later-life functioning.^14,20^

Social class of origin is a parsimonious and policy-relevant summary of childhood conditions in Britain.^21^ Social class origin structures educational opportunities, labour-market entry and trajectories of working life, and it is therefore a plausible determinant of cognitive outcomes in later life.^21,22^ A natural question follows: can educational expansion and working-life social mobility reduce the enduring association between social origin and cognitive outcomes? Much research constructs mobility as an origin-by-destination cross-classification.^23^ While intuitive, this approach can miss the dynamics and heterogeneity of working-life careers. A trajectory-based approach, on the other hand, constructs social mobility as a ‘video’ rather than a snapshot.^24,25^ Prior work using latent class growth models in the 1958 National Child Development Study (British birth cohort) at age 46 indicates that working-life class careers are heterogeneous, that there are a limited number of well-trodden trajectories, and that these trajectories are ordered in advantage.^25,26^ On this basis, working-life social mobility can be implemented as trajectory membership rather than as a single origin-by-destination transition.

Most cognitive ageing studies focus on episodic memory performance over short neuropsychological test intervals (minutes between immediate and delayed recalls).^6,12,26^ These measures are well validated but do not directly assess the integrity of autobiographical retrieval across decades. The logic of life course shaping motivates a longer horizon for episodic memory: whether facts experienced in childhood remain retrievable in early old age.^27^

This study, therefore, analyses two outcomes: (i) standard episodic memory and (ii) validated long-term episodic memory spanning more than half a century. The long-term outcome uses contemporaneous childhood household information recorded by mothers when cohort members were visited at age 11 and re-elicited from cohort members at age 62.^13,28^ Accuracy is coded as a binary indicator: ‘good’ long-term episodic memory is defined as correctly recalling both the number of rooms and the number of people; any mismatch indicates recall error. This is an instance of turning error into substance. The recalled items also underpin an overcrowding measure used in life-course and public health research, and the World Health Organization’s guidelines identify inadequate living space (crowding) as a housing-related health concern.^29^

Using the 1958 British birth cohort, this study examines whether educational expansion and working-life social mobility mitigate the association between social class of origin and episodic memory in early old age. We used derived working-life social mobility for this British cohort as trajectory-based mobility and estimate models separately for women and men.^25,26^ Because cognitive ageing studies are vulnerable to healthy survivor bias, we assess robustness to attrition correction using inverse probability weighting for survival/participation at age 62.^6^

## Methods

This study draws from the 1958 British birth cohort, a prospective study that has followed individuals born in Great Britain during one week in March 1958 across childhood, adolescence and adulthood into early old age. The analysis focuses on the age 62 sweep (2020/21), when cohort members underwent cognitive assessment and provided contemporaneous information on late-life socioeconomic, psychosocial and behavioural information.^26,28^ The analytic sample is restricted to respondents with complete information on outcomes and covariates (N = 4,610; 2,172 men and 2,438 women).

Two cognitive outcomes were analysed. Short-term episodic memory was operationalised as the sum of immediate and delayed word recall (0–20; higher scores indicate better memory). Long-term episodic memory captured the integrity of episodic recall over more than half a century: at age 62 cohort members recalled the number of rooms and the number of people in their home at age 11, and responses were validated against contemporaneous maternal reports. Long-term episodic memory was coded as 1 (‘good’) if both values matched and 0 otherwise.

Social class origin was measured using parental occupation recorded during childhood and coded into a threefold social class category: manual, intermediate and service (derived from Goldthorpe class scheme or ONS-NSSEC class schema).^22^ Working-life social mobility was derived from latent class group trajectories (‘video’ of social class stations rather than snapshot of social class station), then for parsimony classified into a binary indicator distinguishing a predominantly manual class trajectory from non-manual trajectories.^24,25^ All models adjusted for adult socioeconomic attainment (highest education, income, household size), early-life health (time off school for ill health), psychosocial status and social resources (SF-36 mood/anxiety, marital status) and health behaviours (regular exercise, smoking).

Analyses were run separately for women and men. For each outcome-by-sex combination, three nested sub-models were estimated: (1) origin and covariates without mobility and without survivor weighting; (2) adding the mobility indicator; and (3) adding inverse probability weighting to address survival/participation at age 62. Participation is (probit) modelled in terms of prospectively measured parental social class, parental education, childhood ability at age 11, and highest education at 46.^26^ Conceptually, this approach aims to reduce bias arising from the fact that participants at age 62 are on average healthier than those lost to attrition.

Short-term episodic memory was modelled using linear regression. Long-term episodic memory was modelled using probit regression with results presented as average marginal effects for easy interpretation. Modelling was done in Stata version 19.^30^

## Results

### Sample characteristics

The analytic sample comprised 4,610 participants at age 62 (2,172 men; 2,438 women; Table 1). Mean episodic memory was 11.2 (SD 3.1) overall, with higher mean scores among women than men (11.6 vs 10.8). The prevalence of good long-term episodic memory (accurate half-century recall of both rooms and people at age 11) was 36.2% overall (37.1% women; 35.3% men). Social origin was distributed as 58.2% service, 28.8% intermediate and 13.0% manual. The prevalence of a manual work-life class trajectory was 15.4% overall and differed by sex (22.2% men; 9.4% women). More than one in five were educated to degree-level (24.0% women; 23.3% men).

**Table 1.** Sample characteristics at age 62 (mean (SD) or n (%)).

|  | **M (N=2172)** | **F (N=2438)** | **Total (N=4610)** |
| --- | --- | --- | --- |
| Episodic memory | 10.8 (3.0) | 11.6 (3.1) | 11.2 (3.1) |
| **Long term episodic memory, good** |  |  |  |
| No | 1405 (64.7%) | 1534 (62.9%) | 2939 (63.8%) |
| Yes | 767 (35.3%) | 904 (37.1%) | 1671 (36.2%) |
| **Parents class** |  |  |  |
| Manual | 307 (14.1%) | 292 (12.0%) | 599 (13.0%) |
| Intermediate | 623 (28.7%) | 703 (28.9%) | 1326 (28.8%) |
| Service | 1242 (57.2%) | 1441 (59.2%) | 2683 (58.2%) |
| **Social mobility** |  |  |  |
| Non-manual class trajectory | 1690 (77.8%) | 2209 (90.6%) | 3899 (84.6%) |
| Manual class trajectory | 482 (22.2%) | 229 (9.4%) | 711 (15.4%) |
| **Highest qual, short code** |  |  |  |
| None | 161 (7.4%) | 217 (8.9%) | 378 (8.2%) |
| GCSE-A level | 1504 (69.2%) | 1635 (67.1%) | 3139 (68.1%) |
| Degree | 507 (23.3%) | 584 (24.0%) | 1091 (23.7%) |
| **Off school for ill health** |  |  |  |
| 0 | 5 (0.2%) | 6 (0.2%) | 11 (0.2%) |
| Less than 1 week | 1440 (66.3%) | 1467 (60.2%) | 2907 (63.1%) |
| 1 wk to 1 month | 621 (28.6%) | 821 (33.7%) | 1442 (31.3%) |
| 1 to 3 months | 87 (4.0%) | 116 (4.8%) | 203 (4.4%) |
| Over 3 months | 19 (0.9%) | 28 (1.1%) | 47 (1.0%) |
| Total income in GBP thousand | 35.3 (24.8) | 30.7 (23.6) | 32.9 (24.3) |
| Household size | 2.2 (0.8) | 2.1 (0.8) | 2.1 (0.8) |
| SF-36 mood or anxiety | 78.5 (16.9) | 75.4 (18.1) | 76.8 (17.6) |
| **Marital status** |  |  |  |
| Single | 217 (10.0%) | 169 (6.9%) | 386 (8.4%) |
| Married/partnered | 1622 (74.7%) | 1693 (69.4%) | 3315 (71.9%) |
| Widow/divorcee | 333 (15.3%) | 576 (23.6%) | 909 (19.7%) |
| **Exercise regularly** |  |  |  |
| No | 528 (24.3%) | 574 (23.5%) | 1102 (23.9%) |
| Yes | 1644 (75.7%) | 1864 (76.5%) | 3508 (76.1%) |
| **Smoking** |  |  |  |
| non smoker | 1994 (91.8%) | 2232 (91.6%) | 4226 (91.7%) |
| past/current | 178 (8.2%) | 206 (8.4%) | 384 (8.3%) |

### Short-term episodic memory

For women, in all three models, educational attainment was associated with higher episodic memory relative to the reference category of primary education or less (Table 2). GCSE–A level education was associated with approximately 1.10 more words and degree-level education with two more words. Income was positively associated with episodic memory while household size was inversely associated with episodic memory. In the mobility models, a predominantly manual class trajectory was associated with slightly higher episodic memory.

**Table 2.**
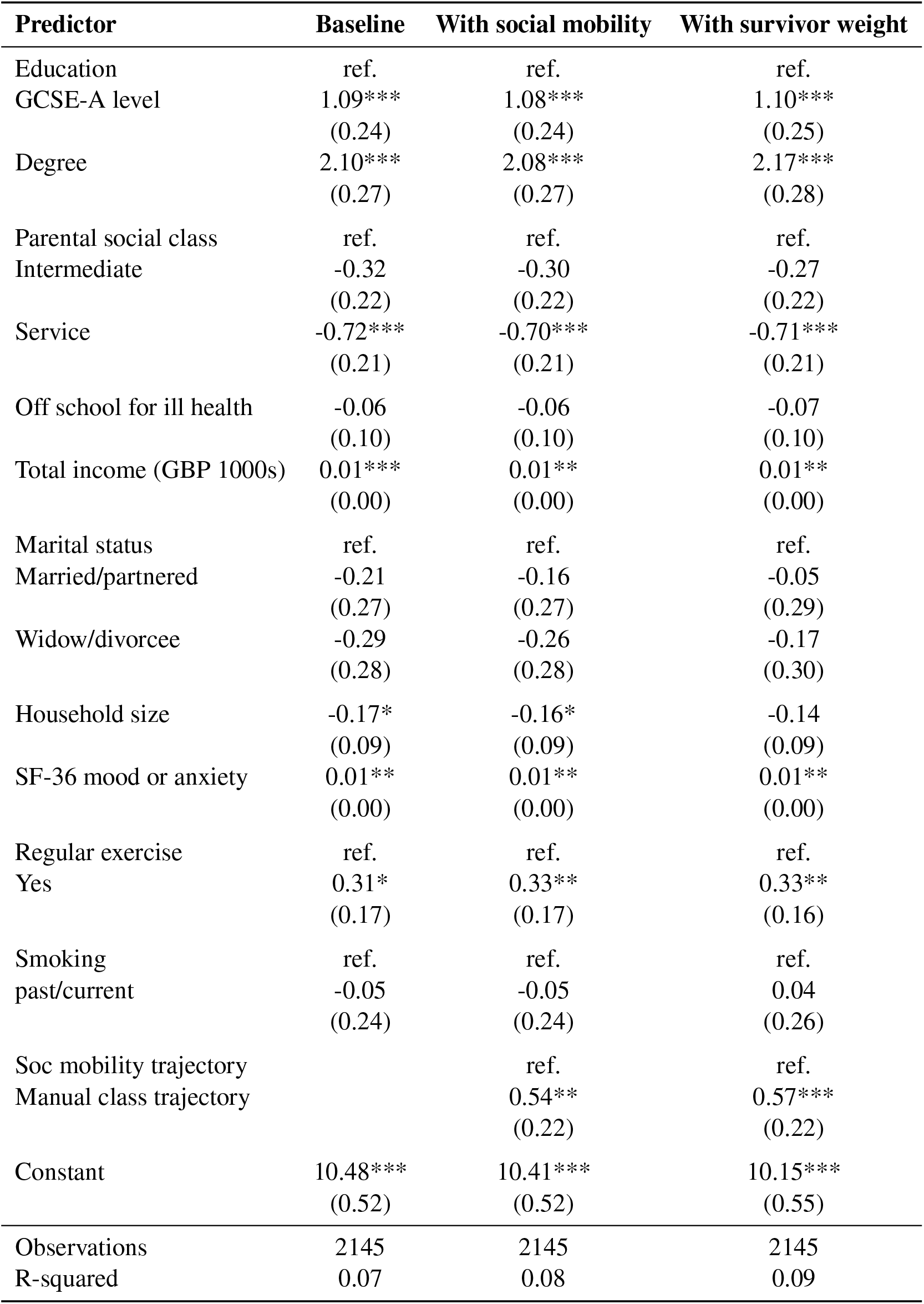
Linear regression for episodic memory among women. Standard errors are shown in parentheses; * *p* < 0.10, ** *p* < 0.05, *** *p* < 0.01.

For men educational attainment was similarly associated with higher episodic memory (Table 3): GCSE–A level education was associated with 1.24 higher points and degree-level education with two more words – both are comparable numbers with women’s. Income was positively associated with episodic memory and so was regular exercise. In mobility models, a predominantly manual class trajectory was associated with higher episodic memory. The service-origin coefficient was statistically significant in the baseline model but attenuated when mobility was included.

**Table 3.** Linear regression for episodic memory among men. Standard errors are shown in parentheses; * *p* < 0.10, ** *p* < 0.05, *** *p* < 0.01.

| Predictor | Baseline | With social mobility | With survivor weight |
| --- | --- | --- | --- |
| Education | ref. | ref. | ref. |
| GCSE-A level | 1.23***<br>(0.27) | 1.19***<br>(0.27) | 1.24***<br>(0.30) |
| Degree | 2.31***<br>(0.30) | 2.18***<br>(0.30) | 2.25***<br>(0.33) |
| Parental social class | ref. | ref. | ref. |
| Intermediate | -0.10<br>(0.22) | 0.03<br>(0.22) | 0.01<br>(0.22) |
| Service | -0.46**<br>(0.20) | -0.23<br>(0.21) | -0.26<br>(0.21) |
| Off school for ill health | 0.07<br>(0.11) | 0.08<br>(0.11) | 0.10<br>(0.12) |
| Total income (GBP 1000s) | 0.01***<br>(0.00) | 0.01***<br>(0.00) | 0.01***<br>(0.00) |
| Marital status | ref. | ref. | ref. |
| Married/partnered | 0.32<br>(0.24) | 0.27<br>(0.24) | 0.23<br>(0.24) |
| Widow/divorcee | 0.12<br>(0.28) | 0.10<br>(0.28) | -0.02<br>(0.28) |
| Household size | -0.11<br>(0.09) | -0.10<br>(0.09) | -0.11<br>(0.09) |
| SF-36 mood or anxiety | 0.01<br>(0.00) | 0.01<br>(0.00) | 0.01<br>(0.00) |
| Regular exercise | ref. | ref. | ref. |
| Yes | 0.47***<br>(0.18) | 0.41**<br>(0.18) | 0.41**<br>(0.19) |
| Smoking | ref. | ref. | ref. |
| past/current | -0.28<br>(0.25) | -0.25<br>(0.25) | -0.16<br>(0.25) |
| Soc mobility trajectory |  | ref. | ref. |
| Manual class trajectory |  | 0.86***<br>(0.17) | 0.88***<br>(0.17) |
| Constant | 8.42***<br>(0.54) | 8.22***<br>(0.54) | 8.20***<br>(0.57) |
| Observations | 1864 | 1864 | 1864 |
| R-squared | 0.08 | 0.10 | 0.10 |

### Long-term episodic memory

For women both GCSE–A level and degree-level education were associated with higher probability of good long-term memory (Table 4). Intermediate origin was associated with a 7% higher probability of good long-term memory compared with manual origin, whereas the service-origin contrast was smaller though not statistically significant. The predominantly manual trajectory indicator was not statistically significant.

**Table 4.** Probit marginal effects for validated long-term episodic memory among women (average marginal effects, SE in parentheses). * *p* < 0.10, ** *p* < 0.05, *** *p* < 0.01.

| Predictor | Baseline | With social mobility | With survivor weight |
| --- | --- | --- | --- |
| Education | ref. | ref. | ref. |
| GCSE-A level | 0.14***<br>(0.04) | 0.14***<br>(0.04) | 0.13***<br>(0.04) |
| Degree | 0.16***<br>(0.04) | 0.16***<br>(0.04) | 0.16***<br>(0.04) |
| Parental social class | ref. | ref. | ref. |
| Intermediate | 0.08**<br>(0.04) | 0.08**<br>(0.04) | 0.07**<br>(0.04) |
| Service | 0.06*<br>(0.03) | 0.06*<br>(0.03) | 0.05<br>(0.03) |
| Off school for ill health | -0.02<br>(0.02) | -0.02<br>(0.02) | -0.02<br>(0.02) |
| Total income (GBP 1000s) | -0.00**<br>(0.00) | -0.00**<br>(0.00) | -0.00**<br>(0.00) |
| Marital status | ref. | ref. | ref. |
| Married/partnered | 0.07*<br>(0.04) | 0.07<br>(0.04) | 0.08*<br>(0.04) |
| Widow/divorcee | 0.10**<br>(0.05) | 0.10**<br>(0.05) | 0.11**<br>(0.05) |
| Household size | 0.01<br>(0.01) | 0.01<br>(0.01) | 0.01<br>(0.01) |
| SF-36 mood or anxiety | 0.00**<br>(0.00) | 0.00**<br>(0.00) | 0.00**<br>(0.00) |
| Regular exercise | ref. | ref. | ref. |
| Yes | -0.02<br>(0.03) | -0.02<br>(0.03) | -0.03<br>(0.03) |
| Smoking | ref. | ref. | ref. |
| past/current | -0.03<br>(0.04) | -0.03<br>(0.04) | -0.02<br>(0.04) |
| Soc mobility trajectory |  | ref. | ref. |
| Manual class trajectory |  | -0.02<br>(0.04) | -0.02<br>(0.04) |
| Observations | 2172 | 2172 | 2172 |

For men educational attainment was associated with higher probability of good long-term episodic memory (Table 5): GCSE–A level education 14% and degree-level education 18%. Origin gradients were evident and aligned with the conceptual order. Compared with manual origin, intermediate origin was associated with 8% higher probability of good long-term memory and service origin with 13%. The predominantly manual trajectory indicator was not statistically significant.

**Table 5.** Probit marginal effects for validated long-term episodic memory among men (average marginal effects, SE in parentheses). * *p* < 0.10, ** *p* < 0.05, *** *p* < 0.01.

| Predictor | Baseline | With social mobility | With survivor weight |
| --- | --- | --- | --- |
| Education | ref. | ref. | ref. |
| GCSE-A level | 0.15***<br>(0.04) | 0.14***<br>(0.04) | 0.14***<br>(0.04) |
| Degree | 0.19***<br>(0.05) | 0.18***<br>(0.05) | 0.18***<br>(0.05) |
| Parental social class | ref. | ref. | ref. |
| Intermediate | 0.07**<br>(0.03) | 0.08**<br>(0.03) | 0.08**<br>(0.03) |
| Service | 0.11***<br>(0.03) | 0.12***<br>(0.03) | 0.13***<br>(0.03) |
| Off school for ill health | -0.01<br>(0.02) | -0.01<br>(0.02) | -0.01<br>(0.02) |
| Total income (GBP 1000s) | -0.00<br>(0.00) | -0.00<br>(0.00) | -0.00<br>(0.00) |
| Marital status | ref. | ref. | ref. |
| Married/partnered | 0.04<br>(0.04) | 0.04<br>(0.04) | 0.04<br>(0.04) |
| Widow/divorcee | -0.04<br>(0.05) | -0.05<br>(0.05) | -0.04<br>(0.05) |
| Household size | -0.03**<br>(0.01) | -0.03**<br>(0.01) | -0.03*<br>(0.01) |
| SF-36 mood or anxiety | 0.00<br>(0.00) | 0.00<br>(0.00) | 0.00<br>(0.00) |
| Regular exercise | ref. | ref. | ref. |
| Yes | -0.00<br>(0.03) | -0.00<br>(0.03) | 0.01<br>(0.03) |
| Smoking | ref. | ref. | ref. |
| past/current | 0.03<br>(0.04) | 0.04<br>(0.04) | 0.04<br>(0.04) |
| Soc mobility trajectory |  | ref. | ref. |
| Manual class trajectory |  | 0.04<br>(0.03) | 0.04<br>(0.03) |
| Observations | 1892 | 1892 | 1892 |

## Discussion and conclusion

Choosing your parents may be the best choice you can make. Despite the post-war expansion of education and the flux of social mobility in Britain,^31^ they have not completely released the grip of social origin on cognition in early old age. The most visible result appears when cognition is measured over a long horizon: validated half-century episodic memory. At 62, only 36% of cohort members retain good long-term episodic memory. And among men, being from the most advantaged origin (service/professional compared to manual class) is associated with 13% higher probability of good memory (a considerable magnitude compared to the 36% prevalence). For men, then, social class reappears with particular clarity when memory is assessed as the ability to retrieve objective childhood facts after five decades. Among women, intermediate origins are associated with a modest advantage over manual origins whereas the service class contrast is smaller and not statistically significant.

The long-term episodic memory findings differ from those of the short-term one in both direction and magnitude. This difference is novel within the episodic memory literature: it warns that conclusions about whether social mobility overcomes social origin depend on how episodic memory is constructed and on the time horizon over which memory is asked to reach. The long-term outcome yields a larger origin gradient than the short-term outcome. For short-term episodic memory, education is the most consistent predictor in both sexes, with degree-level qualifications associated with roughly two extra recalled words relative to primary education or less; GCSE-A level education associated with one extra word. By contrast, the origin coefficients in the short-term models do not consistently show the advantage of service/professional class, particularly in the final model. In both sexes, tracing a manual trajectory is positive, which appears counterintuitive. But these short-term patterns should not be interpreted as a substantive reversal of the origin gradient. Rather, they can be interpreted as conditional associations produced by extensive covariate adjustment and participation at age 62 (healthy survivor bias that remains unaccounted for). Short-term episodic memory is a performance measure taken at a single time point and is therefore sensitive to contemporaneous resources, health behaviours, and psychosocial conditions. Moreover, the model conditioning set includes adult education and income, themselves patterned by origin and plausibly mediate part of the origin–cognition association. Residual healthy survivor bias is an additional interpretation: our cognitive ageing research has shown that attrition is substantial and not ignorable in Britain.^6,8^

In contrast, the long-term episodic memory outcome is both epistemically and conceptually distinct. This is an instance of turning error into substance. It is an integrity measure: it tests whether respondents can retrieve valid childhood facts after five decades. Because the information is checked against mothers’ responses when both mothers and cohort members were visited, recall error and accuracy becomes the performance to be explained rather than a nuisance. This new variable plausibly captures an integrity dimension of memory (accurate retrieval of objective autobiographical facts) more than a performance dimension influenced by immediate neuropsychological test conditions.^31^

Another contribution of this study is the use of intragenerational social mobility as trajectory rather than a cross-classification of origin by destination.^24,25^ Mobility is conceptualised as a “video up to age 46” rather than a snapshot. For parsimony these trajectories are classified into a binary distinction between a predominantly manual trajectory and non-manual one. The results suggest that this long intragenerational social mobility (up to 46 year) fails to remove all the origin effects. For short-term episodic memory, despite weighting, some evidence of unaccounted survivor bias remains – predominantly manual trajectory gives higher episodic memory. But they do not erase the imprint of origin on the integrity of half-century recall – for long-term episodic memory, social origin trumps social mobility.

These results contrast with those from across the Atlantic which tested whether upward mobility overcome childhood disadvantage.^23^ The authors analysed the U.S. Health and Retirement Study (HRS) and reported that upwardly mobile individuals (low childhood socioeconomic status/SES to high adult wealth) were as likely as those with stable high SES to be in the best tertile of several physical health outcomes, concluding that upward mobility can overcome the long arm for those outcomes. To explain this contrast, first note that the childhood SES was derived from recall information which is subject to recall error of unknown magnitude. If the current study is any guide, the error is likely substantial. Moreover, the health systems on both sides of the Atlantic differ considerably, a non-trivial factor.^32,33^

### Strengths and limitations

Three strengths are evident. First, the prospective cohort design provides contemporaneous childhood records, obviating reliance on late-life retrospective reporting for key life course exposures such as social class origin. This design feature avoids confounding between cognitive outcome and measurement error in childhood exposures. Second, the study introduces a novel measure of long-term episodic memory that is validated against objective childhood visits. By transforming recall error from a threat to inference into a cognitive outcome, the study turns error into substance. Importantly, the recalled items, numbers of rooms and people, also underpin an overcrowding measure widely used in life course research.^7,14^ For example, the World Health Organization’s guidelines identify inadequate living space (crowding) as a housing-related health concern, reinforcing that this is a meaningful life course indicator rather than a trivial memory test.^29^ Third, the modelling strategy attempts to account for healthy survivor or attrition bias through weighting for survival to age 62. This approach follows a methodological strand showing that attrition is not ignorable in cognitive ageing studies and that failure to address it can distort inference.^6,8^

The limitations are equally clear. First, cognition is more than memory. The study focuses on episodic memory, and conclusions apply most directly to this cognitive capacity. Other domains such as executive function and processing speed also matter for independent living and retirement decisions.^9,10,15^ Second, age 62 remains early in late life. For this cohort, the state pension age lies beyond 62, so cognition at age 62 captures early old age and the retirement transition rather than oldest-old age.^34,35^ While this enhances relevance for retirement decisions, it also means later-life cognitive decline and dementia-related pathways may not yet be fully expressed. Third, the study does not test biological mechanisms directly. Although the pattern of results is compatible with biological embedding hypotheses, including epigenetic mechanisms for which we demonstrated some support,^14^ the present data do not include direct measures of epigenetic ageing or related biomarkers.

### Mechanisms: epigenetic embedding

The persistence and magnitude of the origin gradient for long-term episodic memory invites a return to biological embedding as a candidate mechanism. Work on the life course shaping of health hypothesis has proposed that childhood poverty can become biologically embedded through epigenetic processes, particularly DNA or RNA methylation, thereby linking childhood poverty to later-life health and functioning.^20,36^ Evidence from the HRS has enabled empirical tests of methylation-based measures of epigenetic ageing in old age and their associations with early-life socioeconomic disadvantage – the same HRS which reported the overcoming of childhood disadvantage by upward social mobility. The HRS epigenetic evidence shows that the childhood poor have more adverse DNA methylation on old age, making them age epigenetically faster.^14^ For the same chronologically age (among the childhood poor and non-poor), the childhood poor’ epigenetic clocks show older ages. While the NCDS 1958 does not furnish epigenetic materials, this mechanism is consistent with the evidence reported here.

### Implication for UN Decade of Healthy Ageing

If adult-stage interventions alone were sufficient to release the hold of social class origin, then policies could focus predominantly on expanding educational attainment and improving labour-market mobility. But the evidence here suggests that such interventions may not be sufficient. Policies targeting childhood conditions remain central to cognitive ageing equity. This conclusion aligns with the framing of the UN Decade of Healthy Ageing (2021–2030), which emphasises the need to add life to years by optimising functional ability and intrinsic capacity and by acting across sectors.^37^ The Decade’s baseline report highlights the importance of environments that support older people’s abilities and of integrated actions across government levels.^38^ A life course reading of the present findings reinforces that responding to population ageing requires earlier intervention than late life: reducing cognitive inequalities in older age requires improving childhood material conditions, not only adult opportunities.

In conclusion, in Britain the post-war education expansion and working-life social mobility have not fully released the hold of social origin on cognition in early old age. The key insight is the divergence between short-term episodic memory and validated long-term episodic memory: when memory is asked to span five decades, the class origin gradient becomes clearer and its magnitude becomes larger. It thus strengthens the case for life course policies that put childhood conditions as core to an effective response to population ageing.

## Data availability

The 1958 National Child Development Study is managed by the UCL Centre for Longitudinal Studies and is freely available for research purposes, accessed via the UK Data Service. Full details and access links can be found at https://cls.ucl.ac.uk/cls-studies/1958-national-child-development-study/.

## Acknowledgement

My work is partly funded by the NIHR Policy Research Unit on Healthy Ageing, University of Manchester. And I thank the Economic and Social Research Council which funds the Centre for Longitudinal Studies (CLS) Resource Centre (ES/W013142/1) and provides core support for the CLS cohort studies. While the CLS Resource Centre makes these data available, CLS does not bear any responsibility for the analysis or interpretation of these data by researchers. The CLS cohorts are only possible due to the commitment and enthusiasm of their participants, I am grateful for their time and contribution.

